# Safety, technical and clinical success of the Aperio Hybrid thrombectomy device in acute ischemic stroke, a prospective post-market clinical follow-up study (HYBRID)

**DOI:** 10.1101/2025.04.01.25324903

**Authors:** Claudia Klüner, Benedikt Sundermann, Catalin George Iacoban, Daniel Behme, Kai Kallenberg, Olaf Wunderlich, Bernd Turowski, Wolfgang Reith, Thomas Liman, Thilo Rusche, Hannes Nordmeyer, Christian Mathys

**Author notes:** Theses authors contributed equally (Claudia Klüner, Benedikt Sundermann). Theses authors contributed equally (Hannes Nordmeyer, Christian Mathys). Corresponding author: PD Dr. med. Christian Mathys Evangelisches Krankenhaus Oldenburg Steinweg 13-17 26122 Oldenburg, Germany.

## Abstract

**Background and Purpose:** Stentretrievers are key devices for endovascular treatment of acute ischemic stroke. Aim of this post market clinical follow-up study was to asses safety and outcomes of interventions using the APERIO® Hybrid/Hybrid^17|21^ thrombectomy device (AHD) in routine clinical use.

**Materials and Methods:** Prospective monitored multicentric national registry of patients with acute intracranial vessel occlusion treated with the AHD in Germany between 11/2020 and 06/2023. Inclusion of patients (n = 173) with low pre-stroke morbidity (mRS <= 2). Assessment of technical recanalization success (mTICI >= 2b), periprocedural symptomatic intracranial hemorrhages (sICH), good clinical outcome (mRS <= 2) at 90 days, and secondary outcomes.

**Results:** Recanalization mTICI ≥ 2b was achieved in 97.1 % of the patients, 84.4 % with the AHD only, including 47.4 % first pass success. Good clinical outcome at 90 days (mRS ≤ 2) was observed in 68.8 %. Good primary safety outcome (no periprocedural sICH) was observed, notably even despite a high rate (79.8 %) of stentretriever oversizing.

**Conclusions:** The AHD is effective and safe. Technical success, primary safety and clinical outcomes surpassed earlier registries on different stentretrievers of the same and earlier generations. However, further studies are warranted to clarify whether these outcomes reflect true advantages of the device, improvements in thrombectomy setups and techniques, or selection biases.

**Key messages:**

- The most important instruments for endovascular stroke treatment, the stent retrievers, differ significantly in design and knowledge about the effects of these differences is limited.
- The APERIO® Hybrid/Hybrid^17|21^ thrombectomy device (AHD) is effective and safe.
- Further studies are warranted to clarify, whether the superior outcomes identified in this registry study reflect true advantages of the AHD.

## 1. Introduction

After numerous studies have demonstrated high efficacy and good safety results for stent retriever-based endovascular stroke treatment, it is recognized as the standard procedure for recanalizing large vessel occlusions in patients with acute ischemic stroke (AIS)^1–7^. However, the standardization of both the procedure and the devices used remains limited.

The main tools used in endovascular stroke treatment, the stentretrievers, differ considerably in design^8^. While clinical outcomes do not vary substantially between conventional designs^9^, innovative design features can have an impact on angiographic outcomes (such as distal embolization rates)^10^. Randomized studies with the purpose to compare different devices are rare^11^, Post market clinical follow-up (PMCF) registries provide additional data on the safety and efficacy of devices in regular clinical use. PMCFs contribute to fulfilling regulatory requirements for continuous clinical surveillance.^12^

Examples of larger registry studies on other stent retrievers are the TREVO Stent-Retriever Acute Stroke (TRACK) registry^13^, the North American Solitaire Acute Stroke (NASA) registry^14^ and the Embotrap Extraction & Clot Evaluation & Lesion Evaluation for NeuroThrombectomy (EXCELLENT) registry.^15^

One aim of the design changes to stentretrievers was to improve their angiographic visibility.^16^ The APERIO® Hybrid/Hybrid^17|21^ thrombectomy device (AHD) is the successor of the Aperio stentretriever (Acandis, Pforzheim, Germany). The newer AHD (CE mark approval in 2019, no FDA approval) incorporates most features of the previous device version such as repeating functional segments of small closed cells and large open cells, aiming to improve integration and retention to the thrombus.^17^ Additionally, it is equipped with embedded radiopaque drawn filled tubing (DFT), which allow full-length visibility of the device under fluoroscopy. Because the additional DFT wires increased the device profile, the initial version of the AHD had to be delivered through a 0.021” microcatheter ^17^. In a second step, a version with a lower profile, compatible with 0.017” microcatheters, was introduced for vessel diameters between 1.0 – 4.0 mm.^18^ Early retrospective multicenter experience indicated a favorable safety profile and optimistic clinical results with the AHD^17–20^, similar to the precursor device.^19^ ^20^ A prospective registry (no monitoring) of anterior circulation strokes treated with the AHD with otherwise broad inclusion criteria confirmed these results.^21^ Additionally, initial positive prospective results of using the AHD in distal intracranial occlusions have been reported (prospective REVISAR registry, congress presentation).^22^ Here we present the results of a prospective, monitored and multicentric PMCF study, collecting comprehensive information on technical and clinical success and safety of the use of the AHD in clinical practice. This study includes both anterior and posterior circulation strokes in patients with relatively low pre-stroke morbidity.

## 2. Methods

### 2.1. General study design

The HYBRID study (clinicaltrials.gov ID: NCT04457479) is a prospective, single arm, multicentric, national PMCF study. The study was approved by the medical ethics committee at the University of Oldenburg (coordinating review board) and subsequently by the local ethics committees at each site. The study was carried out in accordance with the Declaration of Helsinki. Patients or their legal guardians provided informed consent. In cases where consent was not available, e.g. because of death, the need for informed consent to analyze limited initial clinical data (anonymized) was waived. This was done in order to avoid selection biases, mainly due to early mortality after the endovascular procedure.

Originally, eight sites in Germany using or planning to use the AHD were asked to enter each patient treated with AHD into the registry. Seven sites actually included patients between 11/2020 and 06/2023. In the first part of the study, data was collected from routine clinical records in the treatment of patients with AIS using the AHD. The second part of the study consisted of a follow-up by face-to-face or telephone interview planned 90 days after the procedure to obtain information on the clinical outcome and any cerebrovascular events that had occurred in the meantime. Assessment of clinical and image related data was carried out by study-related (neuro-)radiologists locally at each site. All study related data were reviewed by external monitors commissioned by the sponsor.

Pseudonymized data were collected for all patients after informed consent was obtained. In patients with no informed consent available, the follow-up interview after 90 days had to be omitted and anonymized data recording was performed. See Figure 1A for an overview of data acquisition.

**Figure 1A.**
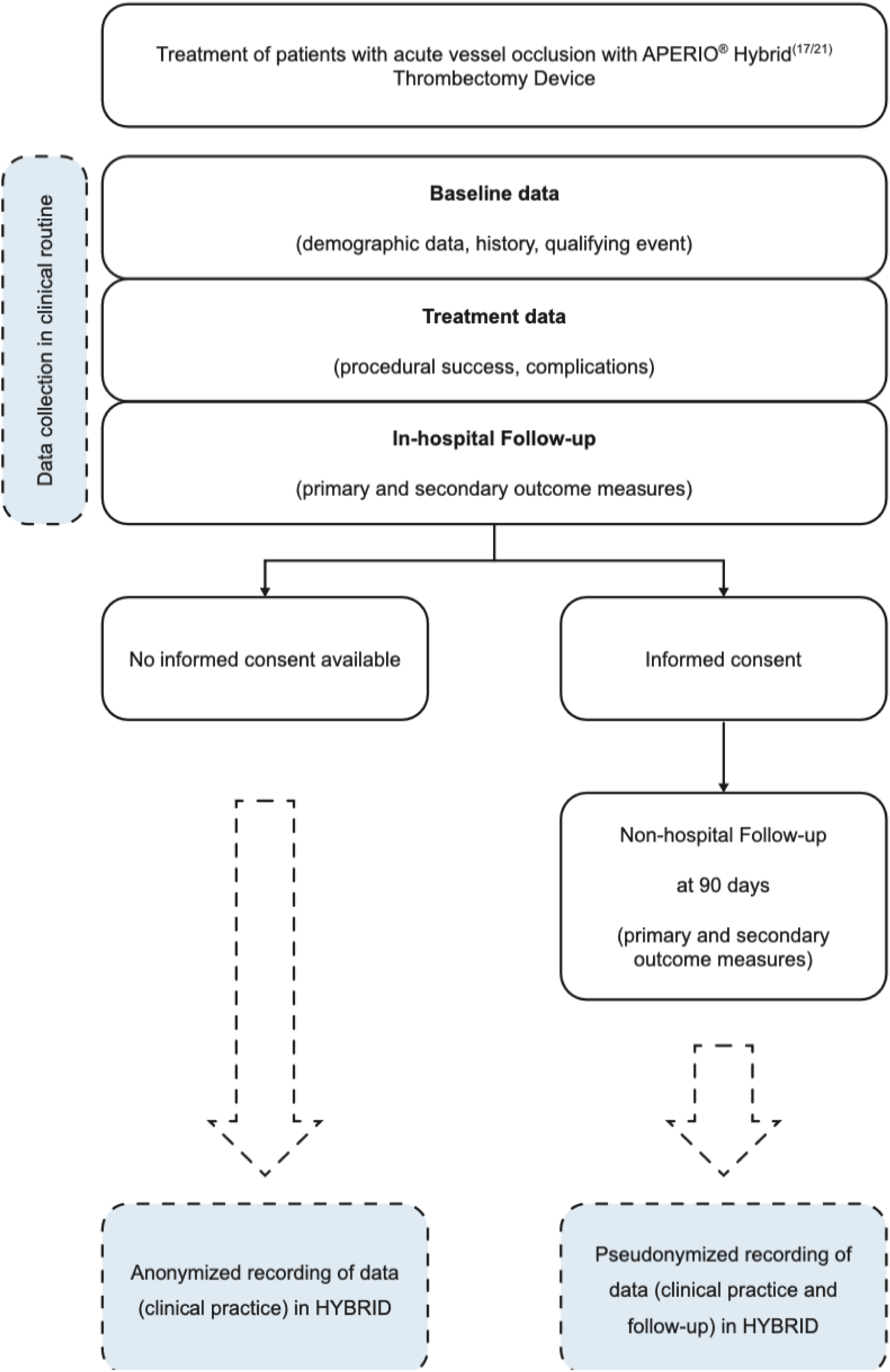
**Data acquisition flowchart.**

### 2.2. In- and exclusion criteria

All patients treated with the AHD (all device sizes) according to the instructions for use (including deviations from the recommended stentretriever sizing) as a result of an AIS (inclusion criterion) were included in the study unless they met one of the exclusion criteria:

- patient age < 18 years^23^, and / or
- pre stroke modified Rankin Scale^24^ ^25^ (mRS) ≥ 3.

### 2.3. Primary outcome measures

The three primary outcomes comprised

- *Technical success*: Modified Thrombolysis in Cerebral Infarction^26^ (mTICI) score ≥ 2b after treatment with AHD,
- *Good clinical outcome at 90 days:* mRS ≤ 2, and
- *Periprocedural symptomatic intracranial hemorrhage (sICH)*. The latter was defined as ICH in the postinterventional (<24 hours) CT associated with worsening of the National Institutes of Health Stroke Scale ^27^ (NIHSS, RRID:SCR_001804) by ≥ 4 points within 24 hours.^28^ ^29^

### 2.4. Secondary outcome measures

Baseline characteristics typical for stroke-related studies were systematically recorded and can be found in Table 1 and in the more detailed supplementary Table S1.

**Tab. 1.**
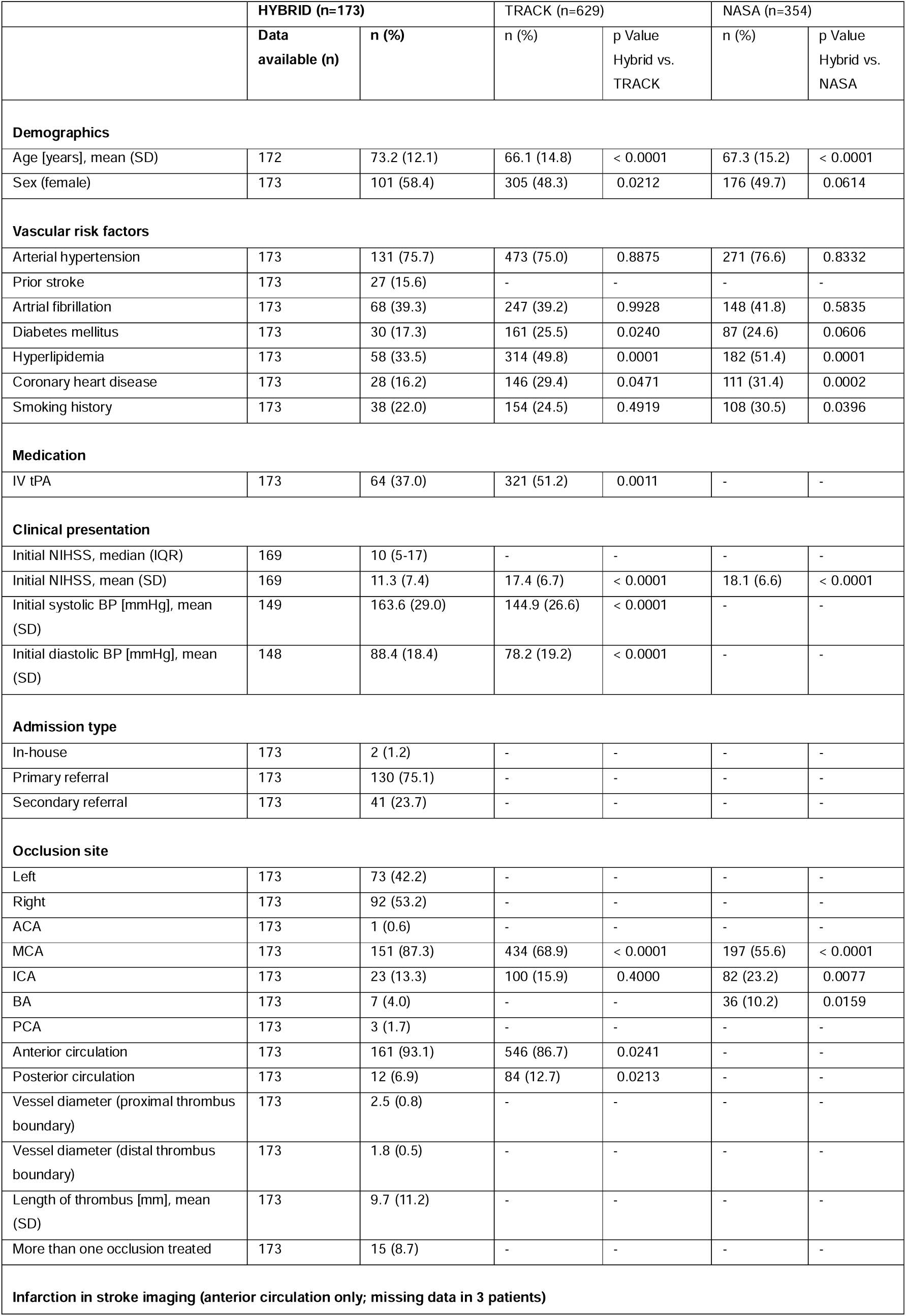
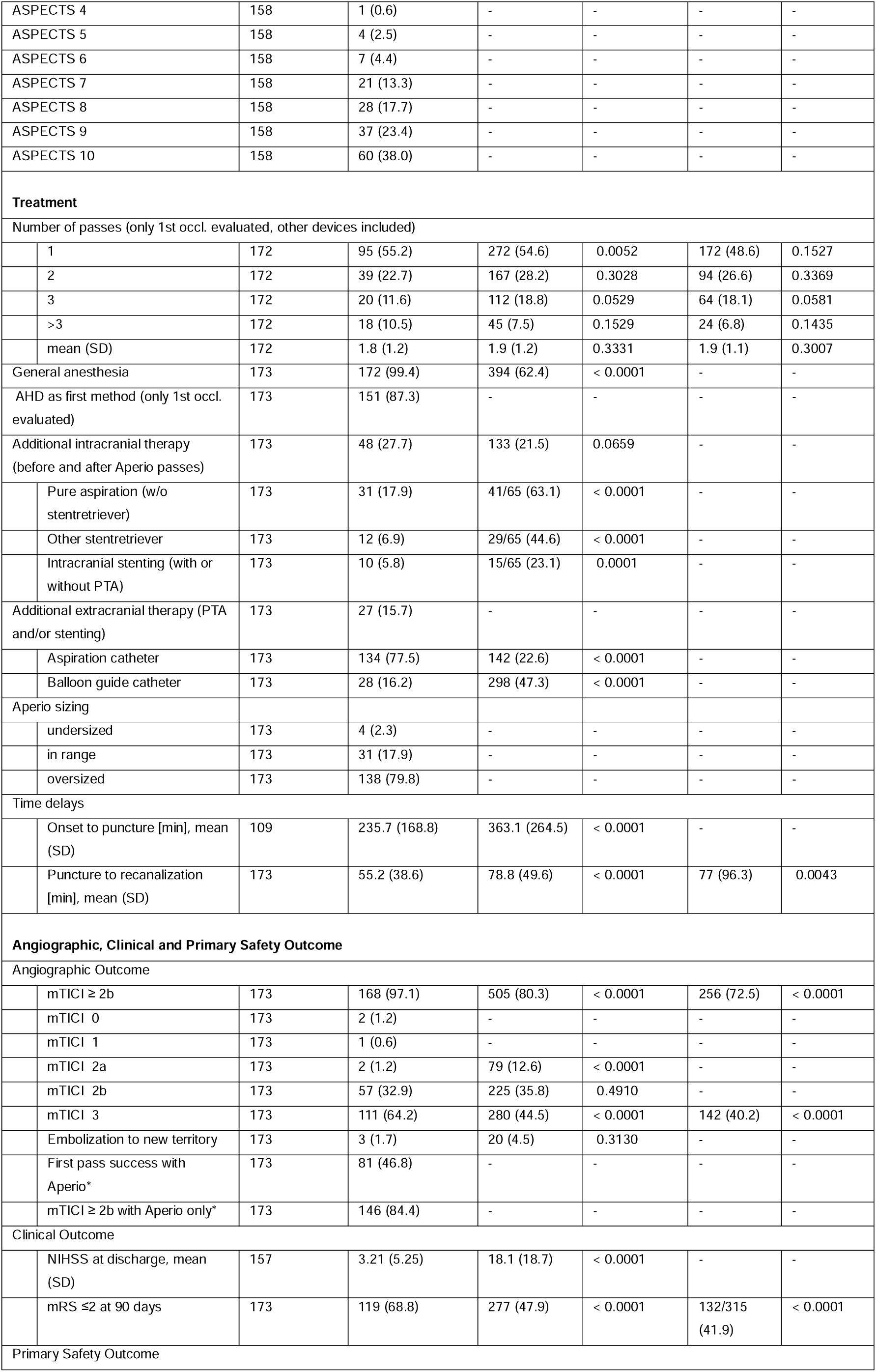
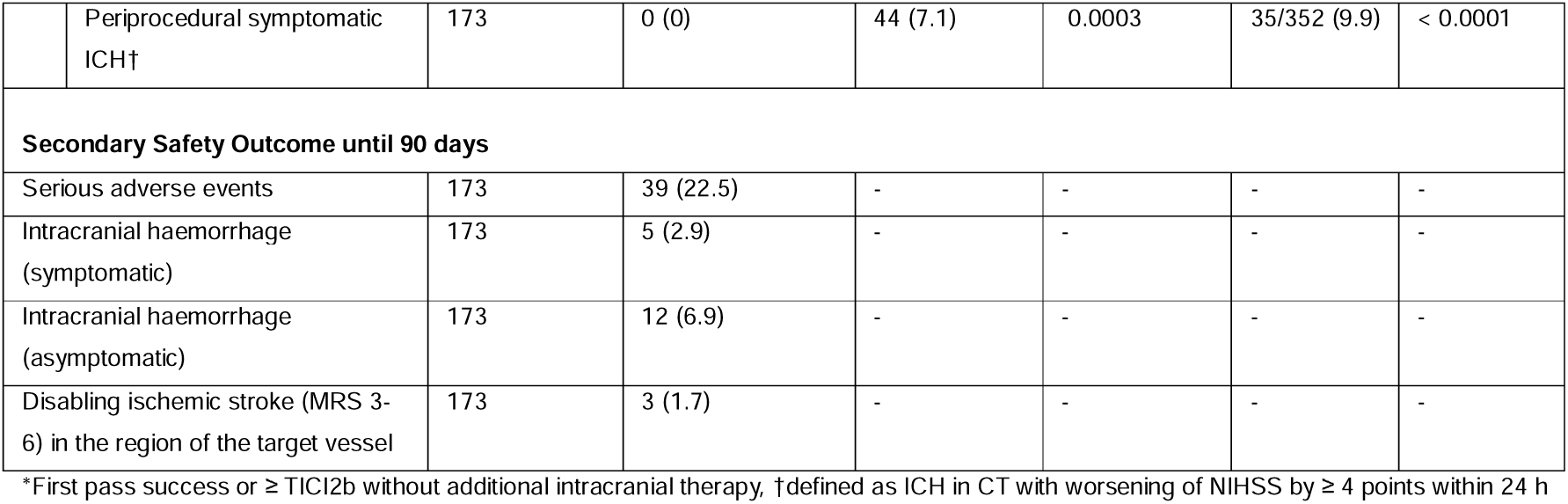
Summary of demographic and clinical characteristics as well as outcomes. The table includes statistical comparison with two earlier, otherwise similar registries (NASA and TRACK) for which table reproduction rights were available. See supplementary Table S1 for a more detailed version and the discussion for a more detailed comparison with other previous studies.

Cerebrovascular events that were collected until hospital discharge comprised: Intracranial hemorrhage (symptomatic / asymptomatic), death, transient ischemic attack (TIA) in the region of the target vessel, non-disabling ischemic stroke (mRS 0-2) in the region of the target vessel, disabling ischemic stroke (mRS 3-6) in the region of the target vessel, TIA outside the region of the target vessel, non-disabling ischemic stroke (mRS 0-2) outside the region of the target vessel and disabling ischemic stroke (mRS 3-6) outside the region of the target vessel.

The following additional cerebrovascular events that occurred between discharge and the follow-up interview at 90 days were also recorded: intracranial hemorrhage (symptomatic / asymptomatic), death, TIA, non-disabling ischemic stroke (mRS 0-2), disabling ischemic stroke (mRS 3-6).

Over the whole period of study participation, the following secondary outcome events were recorded for each patient: all-cause mortality at 90 days, serious adverse events, product-related non-serious and serious adverse events, dissection of the target vessel, occlusion of the target vessel, myocardial infarction, severe extracranial hemorrhage (requiring surgical treatment or transfusion).

### 2.5. Statistical Methods

Although this is a descriptive study, the size of the sample was estimated in such a way that a later metanalytical comparison with other registries or studies on the use of stentretriever systems would be possible with regard to the clinical outcome after 90 days. Based on previous evidence, functional independence (mRS < 3) can roughly be expected in a proportion of 50% of patients after 90 days. Aiming for an error bound of 7.5% (i.e. 15% width of the 95% confidence interval) and expecting a 10% drop-out rate, a sample size of 190 patients was calculated.

The data were processed by an external statistician, commissioned by the sponsor as well as the investigators. Baseline characteristics, technical outcome, clinical outcome and safety related results were analyzed and contrasted with the published results of the TRACK and NASA registries. Student’s t test and F test were used for continuous variables. The χ^2^ and Fisher exact tests were used for categorical variables. Odds ratios and their 95 % confidence intervals were calculated with the formulas given by the MedCalc Odds ratio calculator [MedCalc Software Ltd. Odds ratio calculator. https://www.medcalc.org/calc/odds_ratio.php (Version 23.0.6; accessed October 26, 2024)]. Further statistical analyses were performed using SPSS [IBM SPSS Statistics, Version 29.0.0.0, RRID:SCR_002865] and MATLAB [MathWorks MATLAB Version: 9.12.0.2009381 (R2022a) Update 4, RRID:SCR_001622]. Statistical significance was set at p < 0.05 (not corrected for multiple comparisons).

## 3. Results

### 3.1. Characterization of the clinical sample

A total of 190 patients were included in the HYBRID registry (Figure 1B). Three patients were excluded for reasons that prevented sufficient data collection for the first phase of the study (baseline data, treatment data, hospital follow-up). In this group, one patient withdrew consent and two patients were transferred to another hospital immediately after the procedure. Fourteen patients were excluded, either due to deviation from the study protocol (three patients) or because they were lost to follow-up for reasons other than death (11 patients). The per-protocol set thus contained a total of 173 patients, 156 of whom reached the 90 days follow-up (median: 92, range 80 to 107 days). Seventeen patients died before reaching the 90 days follow-up interview.

**Figure 1B.**
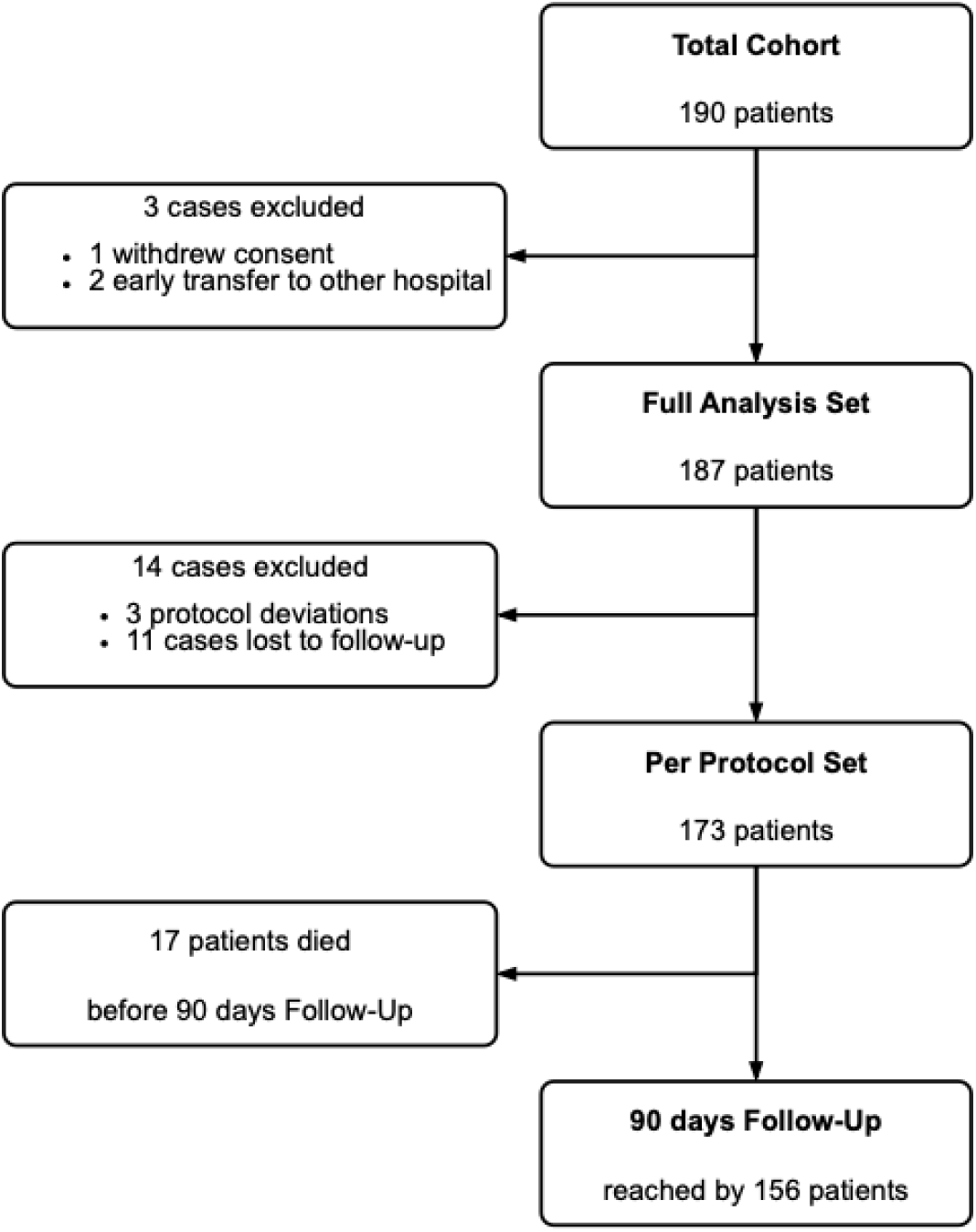
Patient inclusion flowchart. Information on baseline clinical and imaging characterization, type of occlusion and treatment related are presented in Table 1 and in the more detailed supplementary Table S1.

### 3.2. Technical success

Recanalization mTICI ≥ 2b was observed in 97.1% of the patients (see Table 1). A first pass success (mTICI ≥ 2b after a single pass) with the AHD was achieved in 47.4% of the patients. Finally, in 84.4% of the patients technical success (mTICI ≥ 2b) was achieved with the AHD only (no other intracranial therapy necessary like pure aspiration, other stent retriever or intracranial stenting).

Several factors were analyzed for their influence on technical success by calculating the odds ratio and the 95% confidence interval, respectively (Figure 2A). Additional intracranial therapy (pure aspiration, other stent retriever, intracranial stenting and/or angioplasty) was associated with a reduced odds ratio (0.04, 95% CI 0.00-0.35) for technical success.

**Figure 2A.**
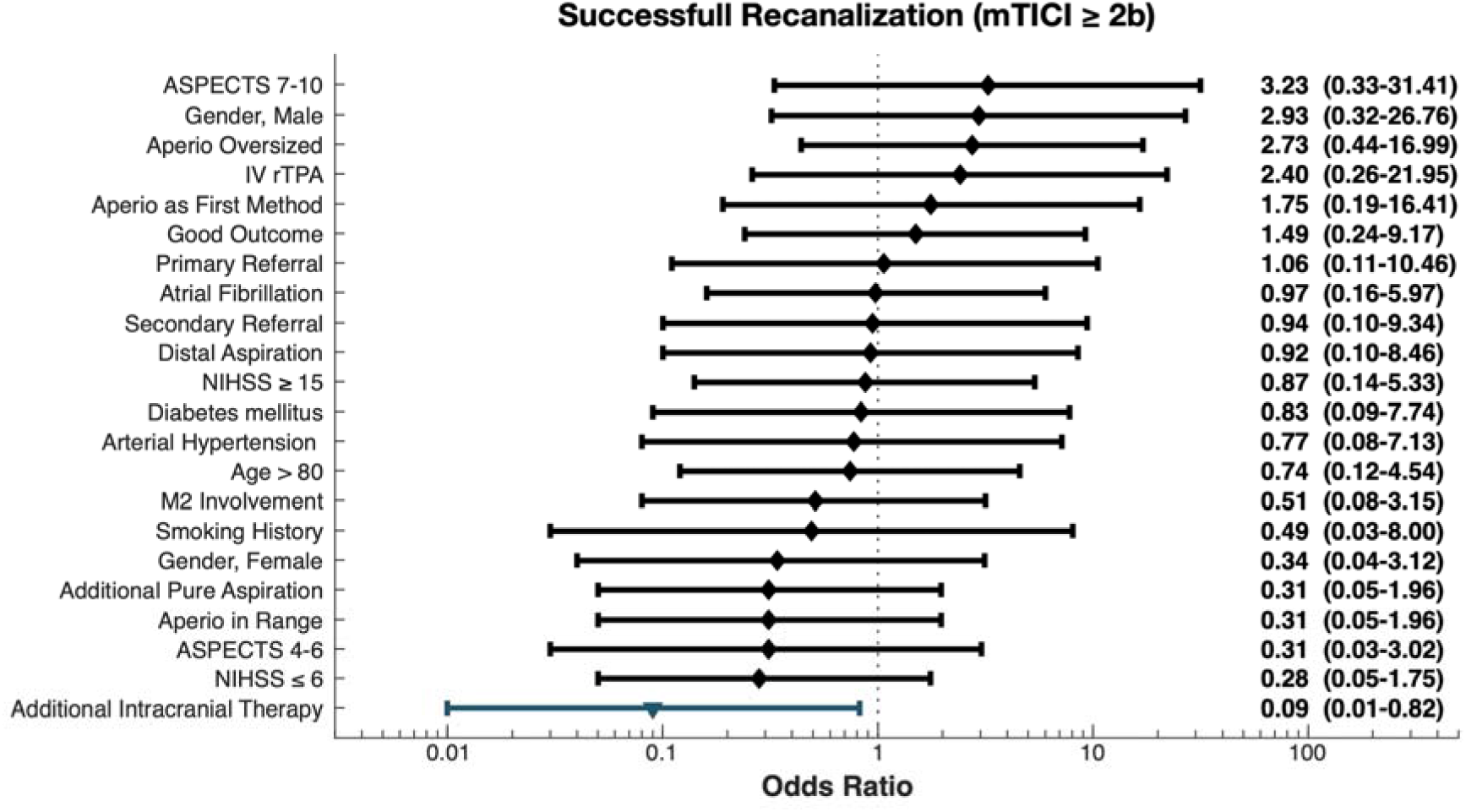
Summary of potential mediators of technical recanalization success. Odds ratios (confidence intervals in parentheses, not corrected for multiple comparisons). Figure design was inspired by the forest plot in the publication of the results of the NASA trial.^14^

**Figure 2B.**
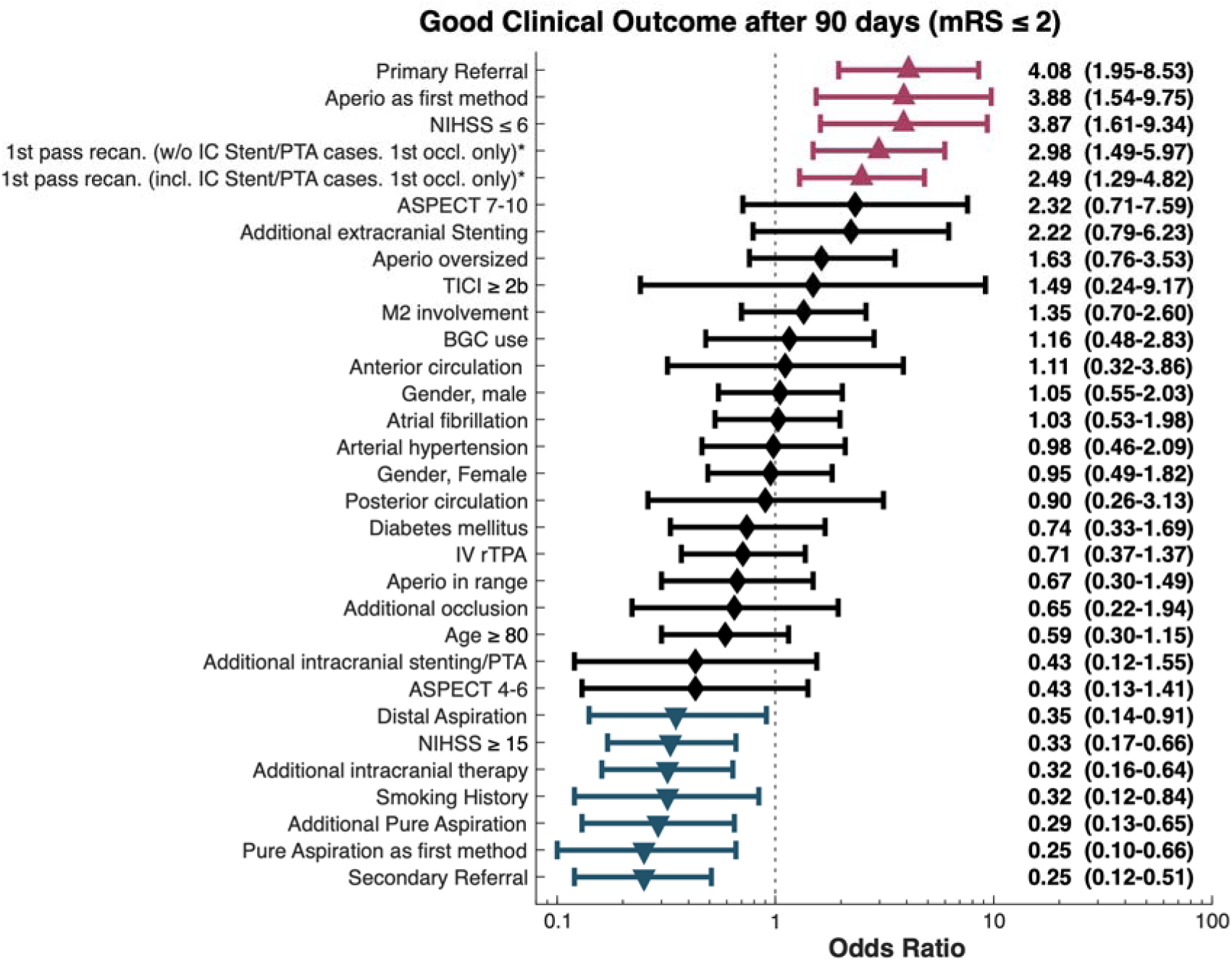
Summary of potential mediators of good clinical outcomes. Odds ratios (confidence intervals in parentheses; red: significant positive association, blue: significant negative association, not corrected for multiple comparisons). Figure design was inspired by the forest plot in the publication of the results of the NASA trial.^14^

### 3.3. Clinical outcome

Mean NIHSS at time of discharge was 3.2 +/- SD 5.3. Good clinical outcome at 90 days (mRS ≤ 2) was observed in 68.8% of patients (see Figure 3 for an overview of the mRS distribution).

**Figure 3.**
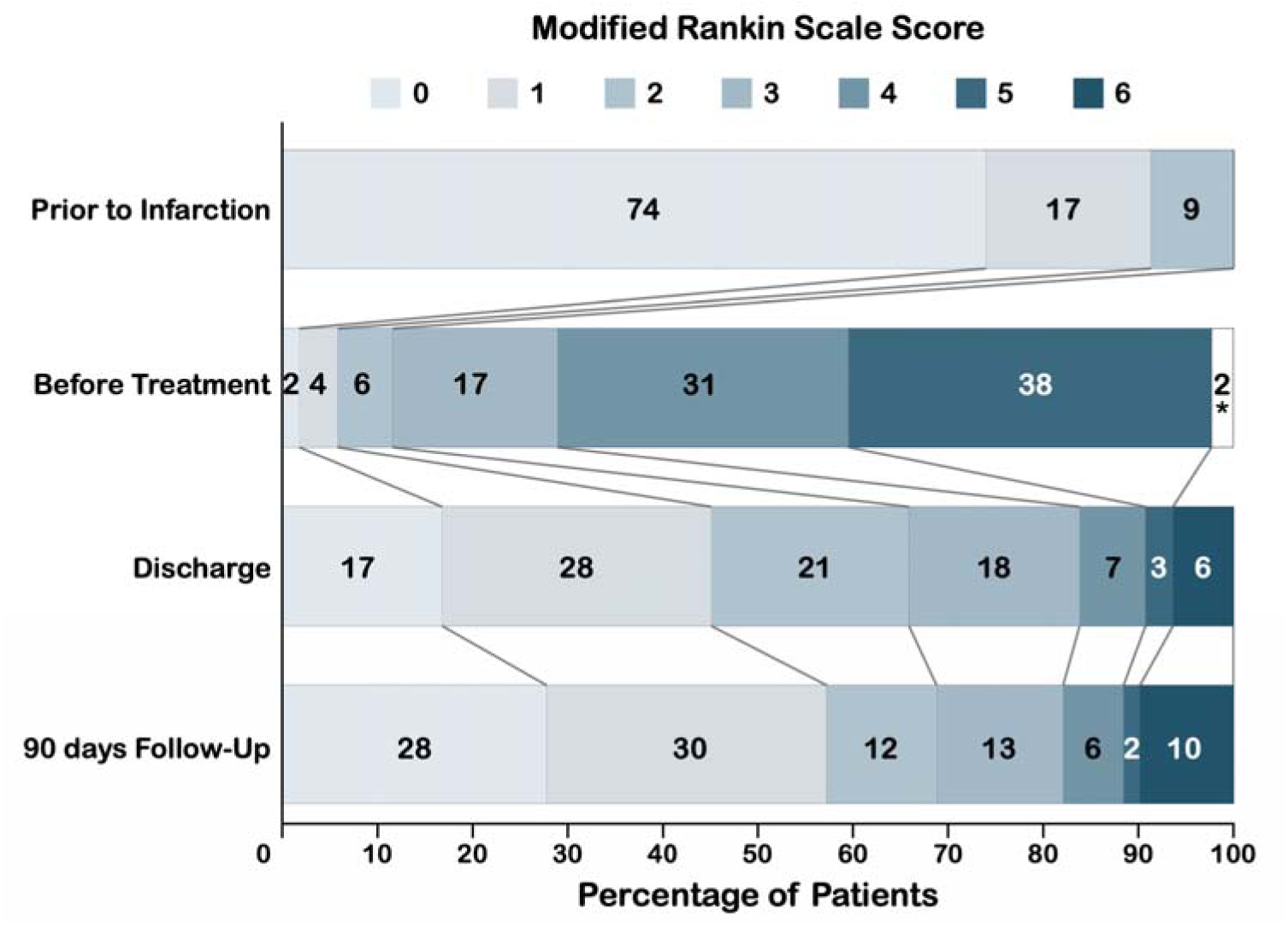
Disability or dependence at different timepoints related to stroke treatment. Percentages do not add to 100 % due to rounding, *missing data.

Several factors were analyzed for their association with good clinical outcome by calculating the odds ratio and the 95% confidence interval, respectively (Figure 2B). Increased odds ratios were found for primary referral (4.08, 95% CI 1.95-8.53), use of the AHD as the first method (3.88, 95% CI 1.54-9.75), NIHSS ≤ 6 (3.87, 95% CI 1.61-9.34), first pass recanalizations without intracranial stent/angioplasty cases (2.98, 95% CI 1.49-5.97) and first pass recanalizations without intracranial stent/angioplasty cases (2.49, 95% CI 1.29-4.82).

Significantly decreased odds ratios were found for distal aspiration (0.35, 95% CI 0.14-0.91), NIHSS ≥ 15 (0.33, 95% CI 0.17-0.66), additional intracranial therapy (0.32, 95% CI 0.16-0.64), smoking history (0.32, 95% CI 0.12-0.84), additional pure aspiration (0.29, 95% CI 0.13-0.65), pure aspiration as the first method (0.25, 95% CI 0.10-0.66) and secondary referral (0.25, 95% CI 0.12-0.51).

### 3.4. Safety outcome

No periprocedural sICH (i.e. < 24 h after the procedure) occurred in HYBRID. However, 17 intracranial hemorrhages which did not meet the criteria of a periprocedural sICH were observed until hospital discharge. No further ICH were reported at 90 days. Five of these were classified as late sICH, while the rest of the bleedings remained clinical silent. In total, 39 serious adverse events were reported until 90 days, of which none was considered device-related. Further secondary safety outcomes are reported in Table 1.

## 4. Discussion

In this prospective, multicentric, externally monitored register including adult patients with AIS predominantly in the anterior circulation, relatively low pre-stroke disability and relatively low early signs of infarction, treated with the AHD, we observed relatively high technical success rates with the AHD only and very high technical success rates in combination with other devices. However, in less than half of the cases mTICI ≥ 2b was achieved as a first pass effect with the AHD. Both, initial safety outcomes and clinical outcomes at 90 days were favorable.

### 4.1. Technical success

The recanalization rate in our registry study was relatively high, with an mTICI≥2b outcome achieved in 97.1 % of cases. This compares favorably to the successful recanalization (mTICI≥2b) rates reported in the HERMES collaboration meta-analysis (71 %)^30^, the data from the STRATIS registry (87.9 %)^31^, the TRACK multicenter Registry (80.3 %)^13^, NASA^14^ (72.5%), ARISE II Study (80.2 %)^32^ and EXCELLENT Registry (94.5 %).^33^ Accordingly, complete recanalizations (mTICI = 3) were observed in 64.2% of HYBRID patients, compared to 44.5% (TRACK^13^) and 40.2% (NASA^14^). This discrepancy may be partially attributed to advancements in device/setup technology since data collection of the five major trials included in the HERMES meta-analysis (prior to 2015) as supported by a longitudinal analysis of MR CLEAN registry data.^34^ Additionally, 99.4 % of patients in our registry study were treated under general anesthesia, a factor that may have contributed to the high recanalization rate.^35^

Additionally, our study identified a positive association between mTICI ≥ 2b recanalization and AHD oversizing (OR 2.73; see Figure 2A). Furthermore, successful recanalization was associated with the selection of AHD as the first-line approach (OR 1.75; see Figure 2A). This size-related finding is consistent with a sub-analysis of the STRATIS Registry, which examined the impact of stent retriever size on angiographic outcomes. Specifically, longer stent retrievers (4 × 40 mm) achieved the highest first-pass effect (FPE) and modified FPE compared to larger-diameter or shorter retrievers.^36^ Similarly, Candel et al. reported that stent retrievers with greater length or nominal diameter yielded higher rates of complete and successful FPE, as well as improved recanalization success, particularly in thrombectomy for acute M1 occlusions.^37^

### 4.2. Clinical outcomes

Overall, the mean NIHSS at discharge in the HYBRID cohort was relatively low (3.2 ± 5.3) compared to TRACK^13^ (18.1 +/- 18.7; see Table 1). A good clinical outcome at 90 days (mRS ≤ 2) was achieved in 68.8% of HYBRID patients, exceeding the outcomes reported in HERMES (46 %)^30^, MR CLEAN registry (37,9 %)^38^, STRATIS (56.5 %)^31^, NASA(42%)^14^, the TREVO registry/TRACK (47.9 %)^13^, EXCELLENT (46.8 %)^33^, and ARISE II (67 %)^32^. In the subgroup of patients with medium and distal vessel occlusions (MeVO/DVO) (n = 45), 77.7% achieved a good clinical outcome at 90 days (mRS ≤ 2), surpassing both the overall HYBRID cohort and the outcomes reported in the DISTAL (56.5 %)^39^ and ESCAPE MeVO trials (54 %)^40^.

### 4.3. Safety outcomes

In our study, no periprocedural symptomatic intracranial hemorrhage (sICH) was reported. In contrast, sICH was observed in 7.1% of patients in the TRACK study^13^, 9.9% in the NASA registry^14^, 1.4% in the STRATIS registry^36^, 1.6% in the EXCELLENT registry^33^, 4.4% in the HERMES collaboration meta-analysis^30^, 2% in the SWIFT cohort^41^, 5.6% in the ARISE II study^32^, and 7% in the TREVO 2 trial.^42^ Notably, no periprocedural sICH occurred in cases of AHD oversizing, which accounted for 79.8% of cases. This indicates that the application of the AHD was safely feasible. The incidence of sICH in our HYBRID cohort was thus considerably lower than in the aforementioned trials and registries. However, this discrepancy may be partly attributable to a potential underestimation of sICH in our study, as early mortality cases without subsequent imaging precluded a definitive distinction between stroke-related and procedure-related events, such as undetected sICH, as the cause of death.

### 4.4. Limitations

Technical success rating is based on the older mTICI definition ^26^ without a 2c category (i.e. near-complete recanalization). More recently, it has been questioned, whether this missing TICI 2c/3 should define technical success as a lower bound rather than mTICI 2b.^43–45^ Additionally, technical success was estimated based on local mTICI ratings, usually by the treating neurointerventionalist, which typically leads to an overestimation of recanalization rates.^46^ ^47^ The lack of an imaging core lab also affects pre-treatment imaging measures such as ASPECTS.^46^

Given the lack of a control group, e.g. with a different stentretriever and the lack of prespecified testable hypotheses limits this report mainly to a descriptive statistical approach, supplemented by post-hoc or exploratory analyses. These exploratory statistical analyses include comparisons with two previous registries (TRACK^13^ and NASA^14^) as surrogates for the state-of-the art for conventional stentretriever designs. Direct comparability is, however, limited by less strict inclusion criteria of those previous studies, mainly regarding pre-stroke disability^13^ ^14^ as well as clinical baseline characteristics and procedural details (see Table 1): In particular, HYBRID patients were older, while cardiovascular risk factors were less frequent. The proportion of patients who received intravenous tissue plasminogen activator (tPA) was lower in HYBRID than in TRACK (51.2%). Initial mean NIHSS was lower in HYBRID (11.3 +/- 7.4) than in both TRACK (17.4 +/- 6.7) and NASA (18.1 +/- 6.6). ICA occlusions were less frequent than in NASA, while MCA occlusions were more frequent in HYBRID than in both TRACK and NASA. Only 6.9 % of patients in HYBRID had posterior circulation occlusions, which was less frequent than in TRACK (12.7%). General anesthesia was used in nearly all patients in HYBRID,while this was the case in only 62.4% of patients in TRACK. The use of balloon guide catheters (BCG) was less common in HYBRID (16.2%) than in TRACK (47.3%), while aspiration catheters were used more frequently in HYBRID. Time delays were shorter in HYBRID compared to TRACK and NASA. Furthermore, NASA adopted the original TICI rather than the mTICI score.^14^

Analysis and interpretation of the potential impact of stentretriever sizing^36^ ^48^, especially the relatively high rate of diameter oversizing in relation to the target vessel diameter are limited by a potential site bias, caused by the treating neurointerventionalists’ sizing preferences. Otherwise, potential site biases were addressed by the HYBRID study’s multicenter design and external monitoring.

The high technical success rate and relatively good clinical outcomes observed in this study compared with previous registries^13^ ^14^ do not necessarily reflect device-specific benefits of the AHD or the other devices and techniques used to treat patients included in this PMCF registry. In addition to possible reasons discussed in the respective sections, they might in part be related to a selection bias owing to the inclusion being restricted to patients with relatively low pre-stroke morbidity. Results can thus not be directly translated to all real-world treatment scenarios for people with AIS resulting from large vessel occlusion.^49^ Selection bias might also pertain to more patients with favorable outcomes providing informed consent and participating in the follow-up interviews. It cannot be fully excluded that some patients with less favorable outcomes, e.g. those transferred to another hospital early after the procedure might have been missed by screening for study inclusion. This was addressed both by the external monitoring and mandate to include limited clinical data from all consecutive patients treated with the AHD at the participating sites.

## 5. Conclusions

The AHD current-generation stentretriever was both effective and safe. Primary safety (no periprocedural sICH even despite a high stentretriever oversizing rate) and clinical outcomes surpassed earlier registries on different stentretrievers of the same and earlier generations. However, it cannot be distinguished, whether this reflects true device characteristics, refined general endovascular stroke therapy setup and techniques or selection biases which are inherent to PMCF studies in general. While a majority of the patients had large vessel occlusions, this prospective registry also identified above average clinical outcomes in medium- and distal vessel occlusion.

## 6. Acknowledgements

Study monitoring was funded by Acandis GmbH, Pforzheim, Germany.

## 7. Declarations

### 7.1. Ethics

The study was carried out in compliance with the Declaration of Helsinki, applicable ICH-GCP / ISO 14155 guidelines and local/national regulations.

### 7.2. Data availability

Due to German data protection regulations and to safeguard subject confidentiality, data on the level of individual participants cannot be made available (no participant consent for data sharing). Data ownership: Acandis GmbH, Pforzheim, Germany.

### 7.3. Competing interests and Funding

Sponsor and source of funding: Acandis GmbH, Pforzheim, Germany. The study protocol was developed by the principal investigator (C.M.) in collaboration with the sponsor. The sponsor had access to the study data.

#### Christian Mathys

consulting and speaker honoraria from Acandis. consulting and lecturing for Siemens on behalf of the employer (Evangelisches Krankenhaus Oldenburg).

#### Hannes Nordmeyer

consulting and speaker honoraria from Acandis.

#### All other authors

nothing to disclose.

### 7.4. Author contributions

**Claudia Klu□ner:** Validation, Formal Analysis, Investigation, Data curation, Writing – review and editing

**Benedikt Sundermann:** Formal Analysis, Investigation, Writing – original draft

**Catalin George Iacoban:** Investigation, Writing – review and editing

**Daniel Behme:** Investigation, Resources, Writing – review and editing

**Kai Kallenberg:** Investigation, Resources, Writing – review and editing

**Olaf Wunderlich:** Investigation, Resources, Writing – review and editing

**Bernd Turowski:** Investigation, Resources, Writing – review and editing

**Wolfgang Reith:** Investigation, Resources, Writing – review and editing

**Thomas Liman:** Investigation, Resources, Writing – review and editing

**Thilo Rusche:** Formal analysis, Writing – review and editing

**Hannes Nordmeyer:** Investigation, Resources, Writing – review and editing

**Christian Mathys:** Conceptualization, Methodology, Validation, Formal Analysis, Resources, Data curation, Writing – original draft, Visualization, Supervision, Project administration, Funding acquisition

**AI use:** ChatGPT was used to cross check with manual literature research during writing of the introduction and discussion sections.

## Notes

### Author Declarations

Medical Ethics Committee of Oldenburg University (and also of participating sites) gave ethical approval for this work.

