## Supplementary material for "Safety, technical and clinical success of the Aperio Hybrid thrombectomy device in acute ischemic stroke, a prospective post-market clinical follow-up study (HYBRID)": Table S1

| | Aperio Hybrid (n=173) | | TRACK (n=629) | | NASA (n=354) | |
|---|---|---|---|---|---|---|
| | Data Available | N(%) | N(%) | p Value Hybrid versus TRACK | N(%) | p Value Hybrid versus NASA |
| Age [years], mean (SD) | 172 | 73.2 (12.1) | 66.1 (14.8) | < 0.0001 | 67.3 (15.2) | < 0.0001 |
| Sex (female) | 173 | 101 (58.4) | 305 (48.3) | 0.0212 | 176 (49.7) | 0.0614 |
| Arterial hypertension | 173 | 131 (75.7) | 473 (75.0) | 0.8875 | 271 (76.6) | 0.8332 |
| Prior stroke | 173 | 27 (15.6) | - | - | - | - |
| Artrial fibrillation | 173 | 68 (39.3) | 247 (39.2) | 0.9928 | 148 (41.8) | 0.5835 |
| Diabetes mellitus | 173 | 30 (17.3) | 161 (25.5) | 0.0240 | 87 (24.6) | 0.0606 |
| Hyperlipidemia | 173 | 58 (33.5) | 314 (49.8) | 0.0001 | 182 (51.4) | 0.0001 |
| Coronary heart disease | 173 | 28 (16.2) | 146 (29.4) | 0.0471 | 111 (31.4) | 0.0002 |
| Smoking history | 173 | 38 (22.0) | 154 (24.5) | 0.4919 | 108 (30.5) | 0.0396 |
| Current smoker | 173 | 27 (15.6) | - | - | - | - |
| Former smoker | 173 | 11 (6.4) | - | - | - | - |
| Antiplatelets | 173 | 45 (26.0) | - | - | - | - |
| ASA | 173 | 42 (24.3) | - | - | - | - |
| Clopidogrel | 173 | 5 (2.9) | - | - | - | - |
| other Antiplatelet | 173 | 1 (0.6) | - | - | - | - |
| Anticoagulation | 173 | 41 (23.7) | - | - | - | - |
| Statins | 173 | 57 (32.9) | - | - | - | - |
| Antihypertensives | 173 | 122 (70.5) | - | - | - | - |
| Antidiabetics | 173 | 14 (8.1) | - | - | - | - |
| IV tPA | 173 | 64 (37.0) | 321 (51.2) | 0.0011 | - | - |
| mRS prior to Infarction | | | - | - | - | - |
| 0 | 173 | 128 (74.0) | - | - | - | - |
| 1 | 173 | 30 (17.3) | - | - | - | - |
| 2 | 173 | 15 (8.7) | - | - | - | - |
| mRS before treatment (missing data in 4 patients) | | | - | - | - | - |
| 0 | 169 | 3 (1.8) | - | - | - | - |
| 1 | 169 | 7 (4.1) | - | - | - | - |
| 2 | 169 | 10 (5.9) | - | - | - | - |
| 3 | 169 | 30 (17.8) | - | - | - | - |
| 4 | 169 | 53 (31.4) | - | - | - | - |
| 5 | 169 | 66 (39.1) | - | - | - | - |
| Initial NIHSS, median (IQR) | 169 | 10 (5-17) | - | - | - | - |
| Initial NIHSS, mean (SD) | 169 | 11.3 (7.4) | 17.4 (6.7) | < 0.0001 | 18.1 (6.6) | < 0.0001 |
| Initial systolic BP [mmHg], mean (SD) | 149 | 163.6 (29.0) | 144.9 (26.6) | < 0.0001 | - | - |
| Initial diastolic BP [mmHg], mean (SD) | 148 | 88.4 (18.4) | 78.2 (19.2) | < 0.0001 | - | - |

| | | | | | | |
|---|---|---|---|---|---|---|
| In-house | 173 | 2 (1.2) | - | - | - | - |
| Primary referral | 173 | 130 (75.1) | - | - | - | - |
| Secondary referral | 173 | 41 (23.7) | - | - | - | - |
| Left | 173 | 73 (42.2) | - | - | - | - |
| Right | 173 | 92 (53.2) | - | - | - | - |
| Midline | 173 | 8 (4.6) | - | - | - | - |
| ACA | 173 | 1 (0.6) | - | - | - | - |
| MCA | 173 | 137 (79.2) | - | - | - | - |
| MCA M2 or more distal | 173 | 46 (26.6%) | - | - | - | - |
| ICA/MCA | 173 | 10 (5.8) | - | - | - | - |
| ICA/MCA/ACA | 173 | 4 (2.3) | - | - | - | - |
| ICA | 173 | 9 (5.2) | - | - | - | - |
| MCA combined | 173 | 151 (87.3) | 434 (68.9) | < 0.0001 | 197 (55.6) | < 0.0001 |
| ICA combined | 173 | 23 (13.3) | 100 (15.9) | 0.4000 | 82 (23.2) | 0.0077 |
| VA (V4) | 173 | 1 (0.6) | - | - | - | - |
| BA | 173 | 7 (4.0) | - | - | 36 (10.2) | 0.0159 |
| BA/PCA | 173 | 1 (0.6) | - | - | - | - |
| PCA | 173 | 3 (1.7) | - | - | - | - |
| Anterior circulation | 173 | 161 (93.1) | 546 (86.7) | 0.0241 | - | - |
| Posterior circulation | 173 | 12 (6.9) | 84 (12.7) | 0.0213 | - | - |
| Vessel diam. (prox. thrombus boundary) | 173 | 2.5 (0.8) | - | - | - | - |
| Vessel diam. (dist. thrombus boundary) | 173 | 1.8 (0.5) | - | - | - | - |
| Length of thrombus [mm], mean (SD) | 173 | 9.7 (11.2) | - | - | - | - |
| More than one occlusion treated | 173 | 15 (8.7) | - | - | - | - |
| **ASPECTS** | | | | | | |
| 4 | 158 | 1 (0.6) | - | - | - | - |
| 5 | 158 | 4 (2.5) | - | - | - | - |
| 6 | 158 | 7 (4.4) | - | - | - | - |
| 7 | 158 | 21 (13.3) | - | - | - | - |
| 8 | 158 | 28 (17.7) | - | - | - | - |
| 9 | 158 | 37 (23.4) | - | - | - | - |
| 10 | 158 | 60 (38.0) | - | - | - | - |
| **Number of passes (only 1st occl. evaluated, other devices included)** | | | | | | |
| 1 | 172 | 95 (55.2) | 272 (54.6) | 0.0052 | 172 (48.6) | 0.1527 |
| 2 | 172 | 39 (22.7) | 167 (28.2) | 0.3028 | 94 (26.6) | 0.3369 |
| 3 | 172 | 20 (11.6) | 112 (18.8) | 0.0529 | 64 (18.1) | 0.0581 |
| >3 | 172 | 18 (10.5) | 45 (7.5) | 0.1529 | 24 (6.8) | 0.1435 |
| median (IQR) | 172 | 1 (1-2) | - | - | 2 (1-2) | - |

| | | | | | | |
|---|---|---|---|---|---|---|
| mean (SD) | 172 | 1.8 (1.2) | 1.9 (1.2) | 0.3331 | 1.9 (1.1) | 0.3007 |
| **General anesthesia** | 173 | 172 (99.4) | 394 (62.4) | < 0.0001 | - | - |
| **First method (only 1st occl. evaluated)** | | | | | | |
| Aperio | 173 | 151 (87.3) | - | - | - | - |
| Pure aspiration | 173 | 20 (11.6) | - | - | - | - |
| Other Stentretriever | 173 | 2 (1.2) | - | - | - | - |
| **Additional intracranial therapy (before and after Aperio passes)** | 173 | 48 (27.7) | 133 (21.5) | 0.0659 | - | - |
| Pure aspiration (w/o stentretriever) | 173 | 31 (17.9) | 41/65 (63.1) | < 0.0001 | - | - |
| Other stentretriever | 173 | 12 (6.9) | 29/65 (44.6) | < 0.0001 | - | - |
| Intracranial stenting (with or without PTA) | 173 | 10 (5.8) | 15/65 (23.1) | 0.0001 | - | - |
| **Additional intracranial therapy (after last Aperio pass)** | 173 | 26 (16.2) | - | - | - | - |
| Pure aspiration (w/o stentretriever) | 173 | 10 (5.8) | - | - | - | - |
| Other stentretriever | 173 | 10 (5.8) | - | - | - | - |
| Intracranial stenting (with or without PTA) | 173 | 10 (5.8) | - | - | - | - |
| **Additional extracranial therapy (PTA and/or stenting)** | 173 | 27 (15.7) | - | - | - | - |
| **Aspiration lumen (only 1st occl. evaluated)** | | | | | | |
| Aspiration catheter | 173 | 134 (77.5) | 142 (22.6) | < 0.0001 | - | - |
| Balloon guide catheter | 173 | 28 (16.2) | 298 (47.3) | < 0.0001 | - | - |
| Non-balloon guide catheter | 173 | 110 (63.6) | - | - | - | - |
| Long sheath | 173 | 9 (5.2) | - | - | - | - |
| **Aspiration method** | | | | | | |
| Pump | 173 | 62 (35.8 ) | - | - | - | - |
| Syringe | 173 | 111 (64.2) | - | - | - | - |
| **Aperio sizing** | | | | | | |
| undersized | 173 | 4 (2.3) | - | - | - | - |
| in range | 173 | 31 (17.9) | - | - | - | - |
| oversized | 173 | 138 (79.8) | - | - | - | - |
| **Time delays** | | | | | | |
| Onset to puncture [min], mean (SD) | 109 | 235.7 (168.8) | 363.1 (264.5) | < 0.0001 | - | - |
| Puncture to recanalization [min], mean (SD) | 173 | 55.2 (38.6) | 78.8 (49.6) | < 0.0001 | 77 (96.3) | 0.0043 |
| **Angiographic Outcome** | | | | | | |
| mTICI ≥ 2b | 173 | 168 (97.1) | 505 (80.3) | < 0.0001 | 256 (72.5) | < 0.0001 |
| mTICI 0 | 173 | 2 (1.2) | - | - | - | - |
| mTICI 1 | 173 | 1 (0.6) | - | - | - | - |
| mTICI 2a | 173 | 2 (1.2) | 79 (12.6) | < 0.0001 | - | - |
| mTICI 2b | 173 | 57 (32.9) | 225 (35.8) | 0.4910 | - | - |
| mTICI 3 | 173 | 111 (64.2) | 280 (44.5) | < 0.0001 | 142 (40.2) | < 0.0001 |
| Embolization to new territory | 173 | 3 (1.7) | 20 (4.5) | 0.3130 | - | - |
| First pass success with Aperio* | 173 | 81 (46.8) | - | - | - | - |
| mTICI ≥ 2b with Aperio only* | 173 | 146 (84.4) | - | - | - | - |
| **Clinical Outcome** | | | | | | |

| | | | | | | |
|---|---|---|---|---|---|---|
| NIHSS at discharge, median (IQR) | 157 | 2 (1-3) | - | - | - | - |
| NIHSS at discharge, mean (SD) | 157 | 3.21 (5.25) | 18.1 (18.7) | < 0.0001 | - | - |
| mRS ≤2 at 90 days | 173 | 119 (68.8) | 277 (47.9) | < 0.0001 | 132/315 (41.9) | < 0.0001 |
| mRS at 90 days 0 | 173 | 48 (27.7) | - | - | - | - |
| mRS at 90 days 1 | 173 | 51 (29.5) | - | - | - | - |
| mRS at 90 days 2 | 173 | 20 (11.6) | - | - | - | - |
| mRS at 90 days 3 | 173 | 23 (13.3) | - | - | - | - |
| mRS at 90 days 4 | 173 | 11 (6.4) | - | - | - | - |
| mRS at 90 days 5 | 173 | 3 (1.7) | - | - | - | - |
| mRS at 90 days 6 (Mortality) | 173 | 17 (9.8) | - | - | - | - |
| **Primary Safety Outcome** | | | | | | |
| Periprocedural symptomatic ICH† | 173 | 0 (0) | 44 (7.1) | 0.0003 | 35/352 (9.9) | < 0.0001 |
| Serious adverse events | 173 | 39 (22.5) | - | - | - | - |
| Device related non-serious adverse events | 173 | 0 (0) | - | - | - | - |
| Dissection of the target vessel | 173 | 0 (0) | - | - | - | - |
| Occlusion of the target vessel | 173 | 0 (0) | - | - | - | - |
| Myocardial infarction | 173 | 1 (0.6) | - | - | - | - |
| Severe extracranial haemorrhage (req. surgery or transfusion) | 173 | 1 (0.6) | - | - | - | - |
| Intracranial haemorrhage (symptomatic / asymptomatic) | 173 | 17 (9.8) | - | - | - | - |
| Intracranial haemorrhage (symptomatic) | 173 | 5 (2.9) | - | - | - | - |
| Intracranial haemorrhage (asymptomatic) | 173 | 12 (6.9) | - | - | - | - |
| TIA in the region of the target vessel | 173 | 0 (0) | - | - | - | - |
| Non-disabling ischemic stroke (MRS 0-2) in the region of the target vessel | 173 | 0 (0) | - | - | - | - |
| Disabling ischemic stroke (MRS 3-6) in the region of the target vessel | 173 | 3 (1.7) | - | - | - | - |
| TIA outside the region of the target vessel | 173 | 1 (0.6) | - | - | - | - |
| Non-disabling ischemic stroke (MRS 0-2) outside the region of the target vessel | 173 | 0 (0) | - | - | - | - |
| Disabling ischemic stroke (MRS 3-6) outside the region of the target vessel | 173 | 2 (1.2) | - | - | - | - |

*First pass success or ≥TICI2b without additional intracranial therapy

†defined as ICH in CT with worsening of NIHSS by ≥ 4 points within 24 h
